# Pre-analytical variables influence zinc measurement in blood samples

**DOI:** 10.1101/2023.01.12.23284491

**Authors:** David W. Killilea, Kathleen Schultz

## Abstract

Zinc deficiency continues to be a major concern for global public health. The zinc status of a target population is typically estimated by measuring circulating zinc levels, but the sampling procedures are not standardized and thus may result in analytical discrepancies. To examine this, we designed a study that controlled most of the technical parameters in order to focus on five pre-analytical variables reported to influence the measurement of zinc in blood samples, including (1) blood draw site (capillary or venous), (2) blood sample matrix (plasma or serum), (3) blood collection tube manufacturer (Becton, Dickinson and Company or Sarstedt AG & Co), (4) blood processing time (0, 4, or 24 hours), and (5) blood holding temperatures (4°C, 20°C, or 37°C). A diverse cohort of 60 healthy adults were recruited to provide sequential capillary and venous blood samples, which were carefully processed under a single chain of custody and measured for zinc content using inductively coupled plasma optical emission spectrometry. When comparing blood draw sites, the mean zinc content of capillary samples was 0.051 mg/L (8%) higher than venous blood from the same donors. When comparing blood sample matrices, the mean zinc content of serum samples was 0.034 mg/L (5%) higher than plasma samples from the same donors. When comparing blood collection tube manufacturer, the mean zinc content from venous blood samples did not differ between venders, but the mean zinc content from BD capillary plasma was 0.036 mg/L (6%) higher than Sarstedt capillary plasma from the same donors. When comparing processing times, the mean zinc content of plasma and serum samples was 5-12% higher in samples processed 4-24 hour after collection. When comparing holding temperatures, the mean zinc content of plasma and serum samples was 0.5-7% higher in samples temporarily held at 20°C or 37°C after collection. Thus even with the same donors and blood draws, significant differences in zinc content were observed with different draw sites, tube types, and processing procedures, demonstrating that key pre-analytic variables can have an impact on zinc measurement, and subsequent classification of zinc status. Minimizing these pre-analytical variables is important for generating best practice guidelines for assessment of zinc status.

## Introduction

With a likely global prevalence over 1 billion, zinc deficiency ranks as one of the top nutrition challenges worldwide, especially in low and middle-income countries [Fischer 2009, Wessells 2012, Stevens 2022]. Inadequate zinc intake can result in increased susceptibility to infectious disease, impaired child growth, and other serious morbidities [Brown 2004, Black 2008, King 2016]. To address zinc deficiency, several international programs monitor and, when necessary, implement nutrition interventions to improve zinc nutrition in populations of concern [de Benoist 2007, Hess 2007, King 2016]. Zinc status is usually determined by sampling the circulating zinc levels, yet there are important technical challenges that should be considered when attempting to measure zinc within blood [Hess 2007, IZiNCG 2007, King 2016]. First, zinc is a common environmental and industrial contaminant, so all supplies that could have contact with the blood should be certified for use with trace metal analysis, or at least tested beforehand for background zinc levels [Reimold 1978, Keyzer 1983, Brown 2004, IZiNCG 2007, Petersen 2017]. This includes phlebotomy supplies, laboratory plasticware, and reagents used in the preparation of the plasma or serum samples within the laboratory. Second, the blood samples must be handled carefully and processed quickly in order to minimize hemolysis, thought to be the most common error in the clinical lab [Gimenez-Marin 2014]. The concentration of zinc in the erythrocytes is more than 10 times greater than the concentration of zinc in plasma or serum levels, so even a small degree of hemolysis can significantly increase the zinc levels in a blood sample [Iyengar 1978; Taylor 1997, Killilea 2017]. The risk of zinc contamination from internal or external sources is further complicated by the fact that circulating zinc levels are tightly maintained within a narrow homeostatic range under physiological conditions [Johnson 1993, King 2016, Hennigar 2018]. Thus, even small differences in zinc measurements can have important effects on determination of zinc status for a population, or evaluation of a nutrition intervention program. The choices that researchers make for supplies and procedure, i.e. pre-analytic variables, are crucial to success for measuring zinc [Plebani 2006, Yin 2015]. Many of these key variables are discussed on the International Zinc Nutrition Consultative Group (IZiNCG) website (*https://www.izincg.org*).

Pre-analytic variables that affect the way blood is collected include the site of blood draw, use of plasma or serum matrix, and type of blood collection tube (BCT). For blood draw site, the conventional approach for assessing zinc uses venous blood obtained by standard phlebotomy [Hess 2007]. Phlebotomy is a well-established procedure that can produce substantial blood volume from the veins, but requires trained technicians and specific sterile supplies, leading to expense and logistics challenges for larger studies. The most common alternative to phlebotomy is the use of a fingerstick, which is a simple procedure requiring low-cost supplies and minimal training, but typically yields just a few drops of blood. Historically, this low volume of blood prevented the widespread use of fingerstick sampling for zinc analysis, but technological improvements have reduced the amount of blood volume needed for measurement so that fingerstick samples are now more feasible for zinc assessment [Petersen 2017, Kyvsgaard 2019]. However, fingerstick sampling provides blood from the capillary circulation, which is not the same as venous blood. At present, there are few reports comparing capillary to venous samples for zinc assessment. For blood matrix, the choices are almost always plasma or serum. Plasma includes all constituents of the blood except cells, but plasma also requires anticoagulant additives that could potentially interact with zinc [Bowen 2014]. Serum is similar to plasma but with the clotting factors removed so that anticoagulation is not necessary, but serum requires time for clotting and results in loss of plasma proteins which may affect zinc homeostasis. While there have been several studies comparing zinc levels in plasma and serum, there is no consensus as to which matrix type is best for mineral assessment, with some favoring plasma [Kasperek 1981, Banfi 2007] and others favoring serum [Yu 2011, Barroso 2018]. A third format variable relates to manufacturer of the selected BCT(s). Becton, Dickinson and Company (BD) has a major market share for BCTs in the US, whereas Sarstedt AG & Co (Sarstedt) is more common in some European countries. To our knowledge, comparisons of different BCT manufacturers for zinc assessment has not received attention previously.

Pre-analytic variables that affect the way blood is prepared for analysis include processing time and holding temperature for collected samples. It is considered best practice that whole blood samples should be processed immediately to plasma or serum for the best chance of high-quality data [IZiNCG 2007]. However, immediate processing is not always possible, particularly in studies in which the laboratory is a substantial distance from the phlebotomy site or when many participants are managed at the same time. Some reports have investigated the impact of processing time on zinc values, but there is no consensus as to which timepoint(s) should be used as a limit for zinc assessment [English 1988, Tamura 1994, Kraus 2016, Chovelon 2018, Leger 2020]. Similarly, it is considered best practice to store whole blood samples in a refrigerated area prior for later processing [IZiNCG 2007]. Again, this may not always be possible, particularly in field studies where access to a cold chain is limited. Some reports have investigated the impact of holding temperature on zinc values, but there is no consensus as to what temperature protocols should be in place for zinc assessment. Moreover, delays in processing and storage at higher temperatures are likely to co-occur in larger studies and in areas with limited infrastructure, so interactions between the two variables should also be examined.

The methods for blood collection and processing can influence the quality of zinc measurements, but it is difficult to isolate the impact that specific pre-analytic variables have on this assessment. Logistical issues, including supply chain choices or procedural steps, are often co-variables that make it challenging to determine which parameters are most consequential for zinc measurements. We sought a more straightforward design that focused on the impact of specific pre-analytic variables on zinc measurement while minimizing the complications from most other technical parameters. Our goal was to determine how blood draw site, blood matrix, tube type, processing time, and holding temperature impact the assessment of circulating zinc levels.

## Methods

### Study Overview

This study on zinc assessment was conducted in parallel with a previously reported study on hemoglobin assessment, with much of the methodology being the same for both reports [Killilea 2022]. Participants completed a demographic questionnaire, underwent anthropometric measurement, and provided sequential blood draws by fingerstick (capillary blood) and then conventional phlebotomy (venous blood). All blood samples were collected at our clinical site and transported to our laboratory under the custody of the study coordinator. Most variables associated with the blood draws and processing were minimized, including the use of the same clinical site, phlebotomist, blood draw procedures, and sample handling procedures (**Table 1**). The CHORI Institutional Review Board approved this study (2019-055), and recruitment was conducted over 7 months.

**Table 1.**
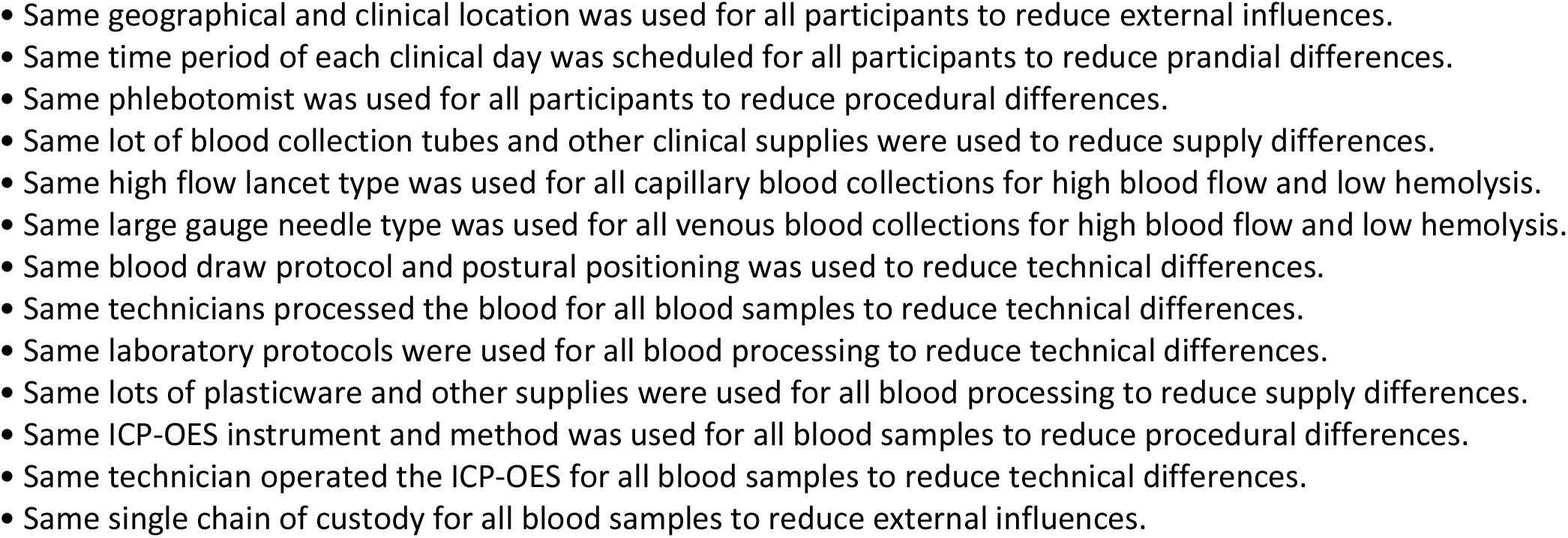
Analytical variables controlled in this study

### Study Participants

A diverse cohort of 60 adults self-described as healthy were recruited from the local community through flyers and word-of-mouth advertising. Inclusion criteria included being 18 years or older, generally healthy with no specified chronic illness or blood disorders. Exclusion criteria included not meeting inclusion criteria or being pregnant or lactating due to known physiological differences in nutrient homeostasis. Participants were asked to avoid consuming vitamin, mineral, and other supplements for 24 hours before their blood draw. Participants were also given a questionnaire asking for self-report of age, gender, and race/ethnicity for use as markers of diversity. Race/ethnicity options were merged to a single choice and used qualitatively since race is a social construct with little scientific value. The cohort recruited for this study were diverse in terms of age, gender, race/ethnicity, and other factors (**S1 Fig and S1 Table**).

### Clinical Procedures

This study was conducted entirely on the campus of Children’s Hospital Oakland Research Institute (CHORI), now part of the University of California San Francisco. All clinical visits were scheduled within the same 2-hour window of the day (10:00AM–12:00PM), as time of day is known to have a strong influence on circulating zinc levels [Hambidge 1989, King 2016]. The clinical procedures began with each participant reviewing and signing the consent form and completing a demographic questionnaire. Height and weight were measured using a Harpenden 602VR stadiometer and Scaletronix ST scale, respectively; these values were used to calculate body mass index (BMI) as mass (kg)/height (m)^2^. The participants then provided non-fasting blood samples beginning with fingerstick(s) then immediately followed by phlebotomy (**S2 Table**). The timing for the blood draws and approximate blood volumes in the BCTs were recorded by the study coordinator. After blood collection, participants were provided juice and snack food while they were observed for recovery. Participants received a gift card after completing the study protocol.

A set of 26 BCTs were prepared and pre-labeled for each participant, including 8 BD plasma tubes (6 ml-size Vacutainer containing dipotassium ethylenediaminetetraacetic acid (K_2_EDTA), product number 368381, lot number 8099632), 8 BD serum tubes (6 ml-size Vacutainer containing clot activator additive, product number 368380, lot number 8099631), 1 Sarstedt plasma tubes (7.5 ml-size Monovette containing K_2_EDTA, product number 01.1605.100, lot number 8031811), 1 Sarstedt serum tube (7.5 ml-size Monovette containing clot activator additive, product number 01.1601.100, lot number 8033711), 4 BD capillary tubes (0.5 ml-size Microtainers containing K_2_EDTA, product number 365974, lot number 8311522), and 4 Sarstedt capillary tubes (0.5 ml-size Microvette containing tripotassium ethylenediaminetetraacetic acid (K_3_EDTA), product number 20.1341.102, lot number 9482211). These BCTs were selected for the goal of comparing zinc levels in blood samples for each participant. For venous blood samples, BD Vacutainer products 368381 and 368380 were used because they are certified for trace metal analysis, rated at a limit of 0.04mg/L zinc according to manufacturer’s instructions. Sarstedt also had venous BCTs designed for trace metal analysis, but only with lithium heparin as the anticoagulant. Since we wished to directly compare BCTs with the same additives, the Sarstedt Monovette products 01.1605.100 and 01.1601.100 were chosen instead even though they were not certified for trace metal analysis. We tested samples from the same lot number of the Sarstedt BCTs and found them to be below detection for zinc content. For capillary blood samples, there are currently no BD products certified for zinc analysis, but the BD Microtainer product 365974 is certified for measuring lead according to manufacturer product information, which may indicate low levels of environmental contaminants. Thus we used the BD 365974 product for capillary blood collection but tested samples from the same lot number and found them to be below detection for zinc content. Similarly, there are currently no Sarstedt products certified for zinc analysis, but the Sarstedt Microvette product 20.1341.102 uses a similar anticoagulant and is also certified for measuring lead according to manufacturer product information. Thus we used the Sarstedt 20.1341.102 product for capillary blood collection but tested samples from the same lot number and found them to be below detection for zinc content.

### Capillary blood collection

To begin the blood collection protocol, each participant was asked to stand to maximize the effects of gravity on blood release from the fingerstick while leaning against a bench for comfort. The participants’ hands were first warmed in a waterbath for 10-20 minutes, dried, and then disinfected using alcohol wipes. Capillary blood was then collected by fingerstick using a high flow lancet (BD product number 366594, lot number V3V51E9) while the participant remained standing. A fingerstick was first applied to the non-dominant hand thumb and then middle (3^rd^) or ring (4^th^) finger if blood volume from the thumb was insufficient. Explanation for the use of the thumb for capillary blood sampling can be found in a previous report [Killilea 2022]. The phlebotomist gently pressed on the base of the hand to start blood flow, but avoided any massaging, squeezing, or milking the thumb or finger [WHO 2010]. The first drop of capillary blood was discarded, then the remaining drops were pooled into a BD Microtainer plasma BCT followed by a Sarstedt Microvette plasma BCT as volume allowed. Capillary blood volume target was 0.5 ml or greater but was stopped short of target volume if the pooled blood from both thumb and finger failed to reach 0.5ml or if the participant requested to end the procedure. If more than 0.5ml was collected, multiple capillary BCTs would be used up to 2ml total volume. Using this fingerstick protocol, 1-2 ml of capillary blood was collected from most participants [Killilea 2022]. Once the capillary blood was collected in all BCTs, tubes were inverted 4-6 times according to manufacturer’s instructions and placed on ice while venous blood collection was completed.

### Venous blood collection

After capillary collection, each participant was moved to a blood collection chair and seated position for phlebotomy [WHO 2010]. Venous blood was then collected by venipuncture in the arm of non-dominant hand, typically at the antecubital vein using a Sarstedt Safety Multifly Needle 21Gx3/4”-size with Multi-Adaptor (product number 85.1638.200, lot number 9050111), as this blood draw system accommodated both BD and Sarstedt BCT types. Venous blood was collected in the following order: Sarstedt Monovette serum BCT, Sarstedt Monovette plasma BCT, BD Vacutainer serum BCTs, and then BD Vacutainer plasma BCTs. Between the BD and Sarstedt BCTs, a discard BCT with no additive was connected to push through a few drops of blood to prevent any contamination when moving to the different BCT manufacturers. Once the venous blood was collected in all BCTs, the tubes were inverted 8-10 times without shaking according to manufacturer’s instructions and held on ice until all samples were taken to the laboratory. Blood samples were transported directly from the clinical site to the laboratory within 10-20 minutes on average and 60 minutes at maximum.

### Laboratory Procedures

#### Processing

Once the samples arrived at the laboratory, the BCTs were quickly sorted and processed. First, 6 of the BD Vacutainers for both plasma and serum were randomly selected for use in the delayed processing experiments, with 2 placed at 4°C, 2 placed at 20°C, and 2 placed at 37°C. After 1 hour, all BCTs at 20°C and 37°C samples were moved to 4°C storage. After 4 hours, 1 BD plasma and 1 BD serum BCT from each of the incubation temperatures (total of 6 BCTs) were randomly selected and processed as described as for ‘time 0’ samples. After 24 hours, the remaining BD plasma and BD serum BCT from each incubation temperature (total of 6 BCTs) were selected and processed as described as for ‘time 0’ samples.

All remaining BCTs were processed immediately as ‘time 0’ samples. For plasma BCTs, the whole blood samples were centrifuged at 800x*g* for 15 minutes at 5°C with no brake to gently separate the plasma from the cellular constituents. For serum BCTs, the whole blood samples were first allowed to clot for 30-60 minutes according to manufacturer product information, before centrifugation at 800x*g* for 15 minutes at 5°C with no brake to gently separate the serum from the cellular constituents. The plasma or serum phases were then transferred to new pre-labeled polypropylene tubes (Perfector Scientific product number 4860, lot number 273381) on ice. The plasma and serum samples were then centrifuged at 4000x*g* for 5 minutes at 5°C with brake to remove any contaminating platelets or erythrocytes. The clarified plasma and serum phases were then aliquoted into multiple pre-labeled polypropylene tubes (Perfector Scientific product number 2840, lot number 292721) for zinc analysis or other uses. All aliquoted samples were stored at −70°C until analysis. Samples from the lot numbers of all plasticware utilized in this study was tested and found them to be below detection for zinc content.

#### Analysis of Hemolyzed Samples

The degree of hemolysis in each blood sample was quantified by measuring the amount of hemoglobin released into the plasma or serum [Drabkins 1935]. Hemoglobin content was determined by modified Drabkin’s assay kit (Sigma-Aldrich product D5941) according to previously published protocol [Killilea 2017]. Each plasma or serum sample was measured in triplicate.

#### Analysis of Zinc Content

Zinc content was determined by inductively coupled plasma optical emission spectrometry (ICP-OES). For measuring zinc in plasma and serum samples, 100 μL volumes were transferred to trace metal tested 15 ml conical tubes (Perfector Scientific, product number 2605), digested in 70% OmniTrace nitric acid (VWR) for 12-16 hours, and then diluted to 5% nitric acid with OmniTrace water (VWR). For testing zinc background in BCTs and plasticware, 3ml of 5% nitric acid was rinsed into the devices and then transferred to trace metal tested 15ml conical tubes. All samples were vortexed 10-30 sec, centrifuged at 4,000x*g* for 10 minutes, and analyzed on a 5100 Synchronous Vertical Dual View ICP-OES (Agilent Technologies). The ICP-OES conditions were set at an RF power of 1.2 kW, plasma flow rate of 12 L/min, nebulizer flow rate of 0.7 L/min, auxiliary flow rate of 1.0 L/min, and pump speed of 12 rpm which delivered sample volume into a Seaspray concentric glass nebulizer and a double-pass glass cyclonic spray chamber. Yttrium (5 mg/L) was added as an internal standard in-line with all samples. Zinc was measured at wavelength 202.548 nm, read time was 10 sec, and the detection range was 0.005–5.000 mg/L. The ICP-OES was calibrated using National Institute of Standards and Technology (NIST) traceable elemental standards (Sigma-Aldrich) and validated using Seronorm Trace Element Levels 1 and 2 standard reference materials (Sero AS); every ICP-OES run with samples from this study passed the Seronorm target range for zinc at the start of each run day. Interassay precision for zinc was determined by taking a pooled plasma sample and a pooled serum sample generated from excess blood from this study and analyzing 30 samples of each for zinc content. The %CV was found to be 2.6% for plasma samples and 4.9% for serum samples, which met the optimal quality specifications for zinc measurement in human plasma or serum [Arnaud 2008]. Additionally, an independent research team tested our laboratory for assessment of zinc concentration in numerous samples including human plasma and serum, and they reported that our facility had excellent precision and accuracy [Hall 2021].

### Data Analysis

Sample size was based on a power calculation to detect significant changes between capillary and venous plasma zinc concentration using data from an unpublished previous pilot study. A minimum enrollment of 50 participants was needed to determine an 8% difference in plasma zinc within subjects between capillary and venous values with alpha = 0.05 and beta = 0.19. The potential for sample attrition due to factors including poor capillary volume production, elevated hemolysis, and abnormal zinc values was estimated to be approximately 15%. Therefore, the targeted enrollment was increased to 60 to increase the chances of having 50 or more blood sample in the final cohort.

All graphing and statistical testing were conducted using Prism 9 (GraphPad Software, Inc). Outlier analysis was conducted using the GraphPad ROUT algorithm with Q=0.1% [Motulsky 2006]. Data normality was assessed with the D’Agostino-Pearson omnibus K2 normality test. For comparisons involving blood draw site, matrix, or BCT type, a Pearson correlation matrix, Bland-Altman analysis, and two-tailed paired t test were used. For comparisons involving processing time and holding temperature for collected blood samples, a two-way ANOVA with Tukey multiple comparison test was used. For all analyses, statistical significance was assigned at p<0.05.

## Results

### Collection of blood samples

Sixty diverse, healthy adults were recruited from Oakland, California to provide blood samples for the comparison of 5 pre-analytic variables (blood draw site, blood matrix, tube type, processing time, and holding temperature) that may influence the measurement of zinc content. Most other technical variables were controlled, including use of the same phlebotomist, blood draw procedures, and processing parameters that commonly vary in larger or field studies (**Table 1**). Each participant provided a capillary blood sample by fingerstick followed immediately by a venous blood sample by venipuncture. Procedural details, including venipuncture location and estimated volume, were recorded for each participant, [Killilea 2022]. The collected blood samples were first tested for elevated hemolysis. Plasma or serum samples with a value of 100 mg/dL hemoglobin or higher were rejected due to the likelihood of zinc contamination from erythrocytes as previously reported [Killilea 2017]. Two samples were identified with hemoglobin levels over the threshold, so those data were excluded from future analysis (**S3 Table**).

### Assessment of zinc status

Plasma zinc was first analyzed from venous blood collected in BD BCTs. Measurements of zinc levels in venous plasma were obtained from all 60 participants, with two separate single values removed from triplicate data after outlier analysis (**S4 Table**). Individual averages of zinc content were normally distributed, ranged between 0.460-0.848 mg/L, and had a mean of 0.647 mg/L (n=60) with a CV of 12.8%. The mean plasma zinc content of the participants was close to the suggested cutoff to define low plasma or serum concentration for morning, non-fasting zinc concentration of 0.65 mg/L as defined by IZiNCG [Brown 2004]. In fact, 32 of the 60 participants would be classified as zinc deficiency with these criteria, so this study had a representation of circulating zinc values from normal to low levels. When separated by sex of participants, the mean zinc content was 0.633 ± 0.070 mg/L for female participants and 0.664 ± 0.095 mg/L for male participants, but this was not significantly different by two-tailed unpaired t test (p=0.156). Differences in mean zinc content based on other demographic data including age, race/ethnicity, and BMI were not determined due to small group numbers.

### Comparison of blood draw site on zinc measurement

A comparison of zinc concentrations in capillary and venous blood samples was conducted from study participants collected in BD BCTs (**Table 2 and Fig 1 A-B**). For capillary plasma measurements, one sample did not have enough volume, one sample was found to have excessive hemolysis, and one sample was identified as outlier, resulting in a total of 57 available plasma samples. For venous plasma measurements, data for 60 plasma samples were available with none found to have insufficient volume, excessive hemolysis, or identified as outliers; however, only venous samples with matching capillary values were compared, resulting in a total of 57 datapoints. Both datasets passed normality testing, so parametric statistics were used. The mean zinc concentration was 0.700 ± 0.094 mg/L for capillary and 0.646 ± 0.084 g/dL for venous, indicating that capillary values were elevated by 0.054 mg/L (8%) in comparison to venous blood values with a sequential blood draw from the same participants. This difference was statistically significant (p<0.0001) using a two-tailed paired t-test. Bland-Altman analysis revealed a similar bias of 0.063 mg/L for capillary zinc values compared to venous zinc values.

**Table 2.** Zinc concentrations differ by three pre-analytic variables.

| Comparisons | mean ± SD (g/dL) | N | difference (g/dL) | p-value |
| --- | --- | --- | --- | --- |
| **Blood draw site (all plasma)** |  |  |  |  |
| venous | $0.646 \pm 0.084$ | 57 | | |
| capillary | $0.700 \pm 0.094$ | 57 | +0.054 capillary | $<0.0001$ |
| **Blood matrix (all venous)** |  |  |  |  |
| plasma | $0.649 \pm 0.083$ | 58 | | |
| serum | $0.679 \pm 0.092$ | 58 | +0.029 serum | $<0.0001$ |
| **Blood collection tube (all venous/plasma)** |  |  |  |  |
| BD | $0.647 \pm 0.083$ | 60 | | |
| Sarstedt | $0.644 \pm 0.083$ | 60 | +0.003 BD | ns |
| **Blood collection tube (all venous/serum)** |  |  |  |  |
| BD | $0.679 \pm 0.092$ | 58 | | |
| Sarstedt | $0.670 \pm 0.089$ | 58 | +0.009 BD | ns |
| **Blood collection tube (all capillary/plasma)** |  |  |  |  |
| BD | $0.688 \pm 0.072$ | 20 | | |
| Sarstedt | $0.652 \pm 0.084$ | 20 | +0.036 BD | $p<0.0001$ |

Mean zinc content is shown for specific pre-analytical variables. Each comparison lists the number of samples measured (N), difference in measured zinc content, and p-value if significant. Comparisons that are not statistically different are identified with ‘ns.’

**Figure 1.**
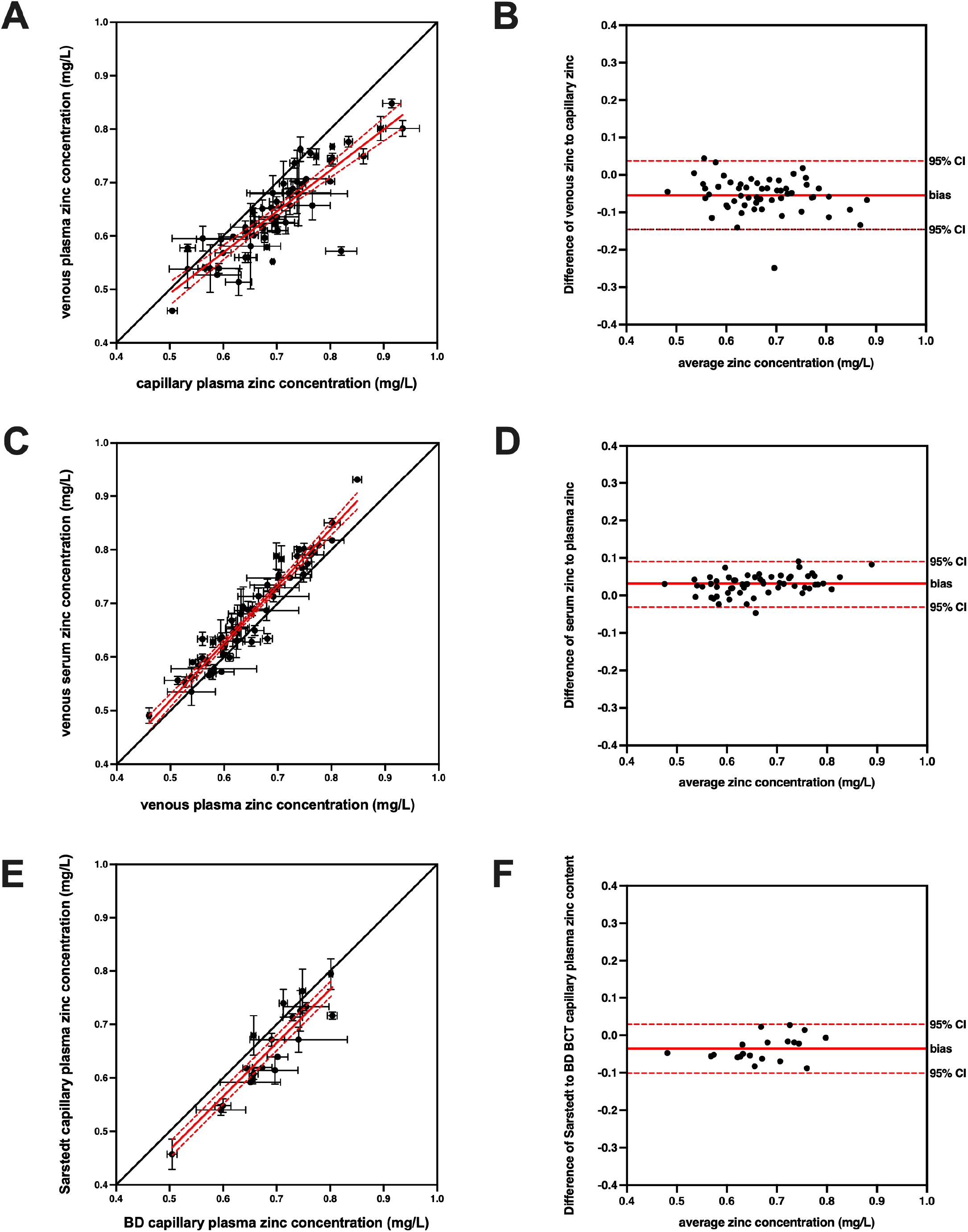
Zinc concentrations differ depending on blood draw site, blood matrix type, and blood collection tube (BCT) manufacturer. Correlation plots comparing circulating zinc values from **(A)** capillary plasma and venous plasma (using only BD BCTs), **(C)** venous plasma and venous serum (using only BD BCTs), and **(E)** capillary plasma from BD and Sarstedt BCTs are shown. Each circle represents the zinc level (mean ± SD, *n*=1-3) for an individual participant, with linear regression and 95% confidence interval indicated by a solid red and dotted red line, respectively. The line of concordance is shown as a solid black line for comparison. Bland-Altman plots of these same data revealed a bias of **(B)** 0.054 ± 0.047 mg/L for capillary over venous plasma samples, **(D)** 0.029 ± 0.025 mg/L for serum over plasma venous samples, and **(F)** 0.036 ± 0.033 mg/L for BD over Sarstedt BCTs for capillary plasma. Each circle represents the zinc level for an individual participant, with average distance and 95% confidence interval indicated by a solid red and dotted red line, respectively.

### Comparison of blood draw matrix on zinc measurement

A comparison of zinc concentrations in plasma or serum was conducted in venous blood from study participants collected in BD BCTs (**Table 2 and Fig 1 C-D**). For venous serum measurements, one sample did not have enough volume and one sample was found to have excessive hemolysis, resulting in 58 available serum samples. For venous plasma measurements, data for 60 plasma samples were available with none found to have insufficient volume, excessive hemolysis, or identified as outliers; however, only plasma samples with matching serum values were compared, resulting in a total of 58 datapoints. Both datasets passed normality testing, so parametric statistics were used. The mean zinc concentration was 0.679 ± 0.092 mg/L for serum and 0.649 ± 0.083 g/dL for plasma, indicating that serum values were elevated by 0.029 mg/L (5%) in a sequential blood draw from the same participants. This difference was statistically significant (p<0.0001) using a two-tailed paired t-test. Bland-Altman analysis indicating a similar bias of 0.029 mg/L for serum zinc values compared to plasma zinc values.

### Comparison of BCT manufacturer on zinc measurement

A comparison of zinc concentrations in BCTs produced by BD or Sarstedt was conducted with venous blood plasma from study participants (**Table 2 and S2 Fig**). For both BD and Sarstedt measurements, data for 60 plasma samples were available with none found to have insufficient volume, excessive hemolysis, or identified as outliers. Both datasets passed normality testing, so parametric statistics were used. The mean zinc concentration was 0.647 ± 0.083 mg/L for BD and 0.644 ± 0.083 g/dL for Sarstedt. This difference was not statistically significant (p=0.4694) using a two-tailed paired t-test.

A comparison of zinc concentrations in BCTs from BD or Sarstedt was conducted with venous blood serum from study participants (**Table 2 and S2 Fig**). For BD serum measurements, one sample did not have enough volume and one sample was found to have excessive hemolysis, resulting in 58 available BD serum samples. For Sarstedt serum measurements, data for 60 plasma samples were available with none found to have insufficient volume, excessive hemolysis, or identified as outliers; however, only samples with matching BD serum values were compared, resulting in a total of 58 datapoints. Both datasets passed normality testing, so parametric statistics were used. The mean zinc concentration was 0.679 ± 0.092 mg/L for BD and 0.670 ± 0.089 g/dL for Sarstedt. This difference was not statistically significant (p=0.0846) using a two-tailed paired t-test.

A comparison of zinc concentrations in BCTs from BD or Sarstedt was conducted with capillary blood plasma from study participants (**Table 2 and Fig 1 E-F**). For Sarstedt capillary measurements, 40 samples did not have enough volume resulting in 20 available Sarstedt capillary plasma samples. For BD capillary measurements, one sample did not have enough volume, one sample was found to have excessive hemolysis, and one sample was identified as outlier, resulting in 57 available BD plasma samples; however, only samples with matching Sarstedt values were compared resulting in a total of 20 datapoints. Both datasets passed normality testing, so parametric statistics were used. The mean zinc concentration was 0.688 ± 0.072 mg/L for BD and 0.652 ± 0.084 g/dL for Sarstedt, indicating that BD capillary plasma zinc values were elevated by 0.036 mg/L (6%) in a sequential blood draw compared to Sarstedt capillary plasma from the same participants. This difference was statistically significant (p<0.0001) using a two-tailed paired t-test. Bland-Altman analysis revealed a similar bias of 0.036 mg/L for BD capillary plasma zinc values compared to Sarstedt capillary plasma zinc values.

### Comparison of processing time and holding temperature on zinc measurement

The effects of delay time and holding temperature on zinc measurements were tested in venous plasma from study participants collected in BD BCTs (**Fig 2 A-B**). Whole blood samples were processed immediately or after a delay of 4 or 24 hours while held at 4°C, 20°C, or 37°C for 1 hour. For plasma samples at time 0-hour, data for 60 plasma samples were available with none found to have insufficient volume, excessive hemolysis, or identified as outliers. For plasma samples at time 4-hour at all temperature settings, three samples were found to have insufficient volume, none had excessive hemolysis, and none were identified as outliers, resulting in 57 available plasma samples. For plasma samples at time 24-hour at all temperature settings, four samples were found to have insufficient volume, none had excessive hemolysis, and none were identified as outliers, resulting in 56 available plasma samples. Only samples with matching values at all temperatures were compared resulting in a total of 56 datapoints. Zinc concentrations for the 4-hour and 24-hour timepoints were all greater than the time 0-hour zinc concentration, ranging from 5-12% higher. Two-way ANOVA of this data indicated that both temperature (p=0.0007) and processing delay (p<0.0001) were a significant variation, but without significant interaction (p=0.1007).

**Figure 2.**
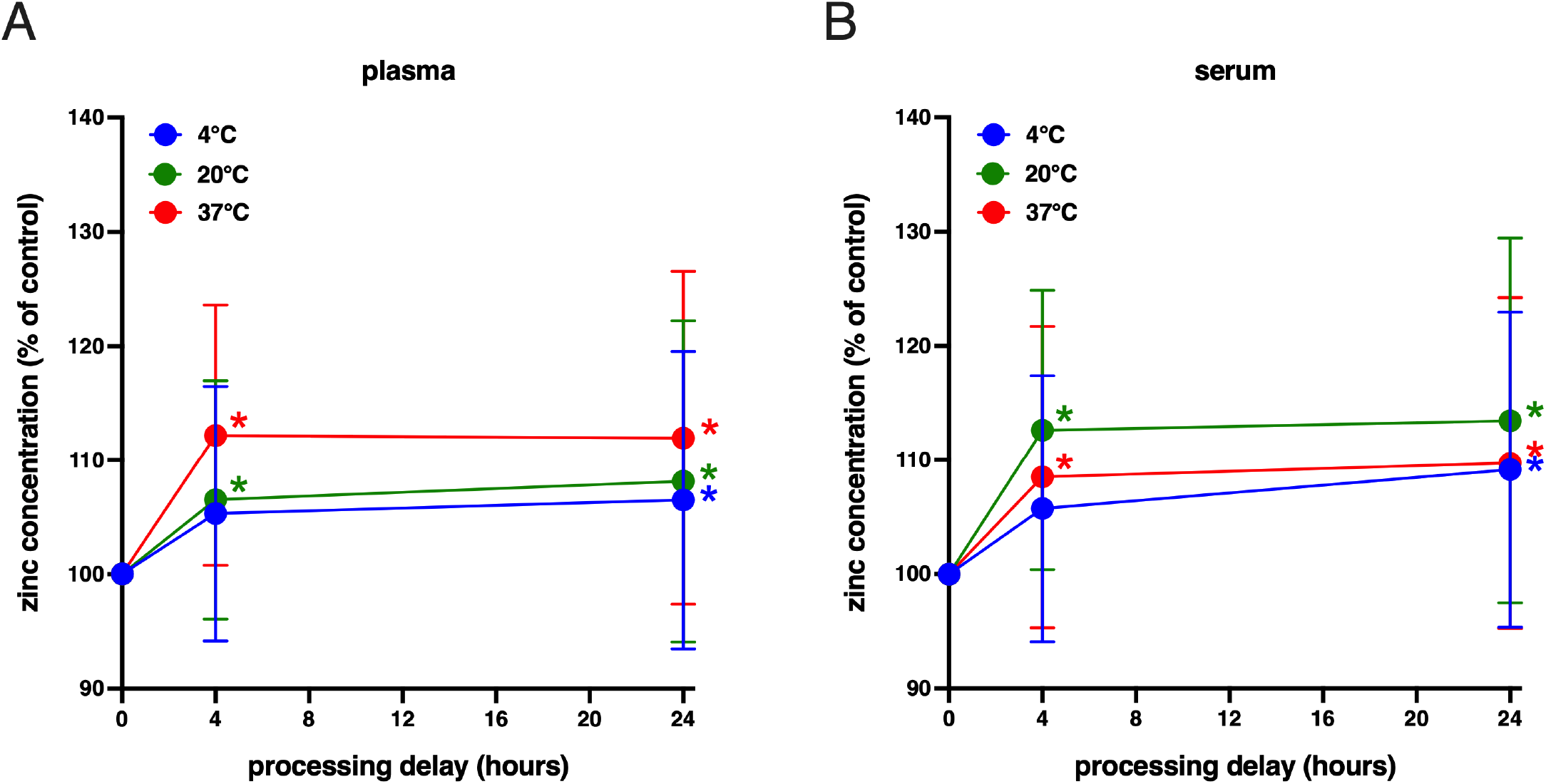
Zinc concentrations differ depending on blood processing time and holding temperature. Timecourse of mean circulating zinc values from blood samples held at 4°C (blue circles), 20°C (green circles), or 37°C (red circles) for up to 24 hours in **(A)** venous plasma and **(B)** venous serum (using BD only BCTs) are shown. Each circle represents the average zinc level (mean ± SD, *n*=55-58) normalized to the ‘time 0’ value for each group of time and temperature condition fit to a point-to-point function. Statistical significance compared to ‘time 0’ of each group is indicated by asterisk of same group color.

The effects of delay time and holding temperature on zinc measurements were also tested in venous serum from study participants collected in BD BCTs (**Fig 2 C-D**). Whole blood samples were processed immediately or after a delay of 4 or 24 hours while held at 4°C, 20°C, or 37°C for 1 hour. For serum samples at time 0-hour, one sample did not have enough volume, one sample was found to have excessive hemolysis, and none were identified as outliers, resulting in 58 available plasma samples. For serum samples at time 4-hour at all temperature settings, no samples were found to have insufficient volume, none had excessive hemolysis, and none were identified as outliers. For serum samples at time 24-hour at all temperature settings, no samples were found to have insufficient volume, none had excessive hemolysis, and none were identified as outliers. Only samples with matching values at all temperatures were compared, resulting in a total of 58 datapoints. Zinc concentrations for the 4-hour and 24-hour timepoints were all greater than the time 0-hour zinc concentration, ranging from 0.5-7% higher. Two-way ANOVA of this data indicated that both temperature (p=0.0061) and processing delay (p<0.0001) were a significant variation, but without significant interaction (p=0.1967).

## Discussion

After two decades of measuring zinc levels in plasma and serum samples for a wide variety of programs, our laboratory has received numerous inquiries about best practices in zinc assessment, most often about blood draw procedures, tube types, or processing concerns. While some of these topics are addressed within the literature on zinc assessment, the available reports are often limited in scope and sometimes conflicting in results. One reason for the lack of consensus may be due to uncontrolled technical parameters that make it difficult to isolate specific pre-analytical variables. The impetus for this study was to focus on specific pre-analytical variables that may impact zinc measurement in order to support best practice guidelines for future assessment of zinc status.

Blood samples from a diverse group of healthy adults were collected and processed under a single chain of custody and highly controlled conditions to test 5 key pre-analytical variables. The mean zinc concentration for venous plasma collected in BD BCTs was 0.647 mg/L, which is near the recommended cutoff of 0.65 mg/L for adequate zinc concentration in adults using morning, non-fasting blood samples [IZiNCG 2007]. Based on this cutoff, a sizeable number of participants in this study would be categorized as mildly zinc deficient despite identifying as healthy. We did not attempt to examine the reasons for the mild zinc deficiency because the primary purpose of this study was only to compare the zinc levels within the same participants and blood draw. In fact, it was helpful to have both zinc sufficient and zinc deficient individuals, as it added to our evaluation of how pre-analytical variables could affect zinc values over a range of circulating zinc concentrations.

The first pre-analytical variable tested was different blood draw sites, specifically the differences in zinc concentration between capillary and venous blood. It is known that capillary and venous blood have different compositions, but the blood matrices are considered similar enough that they have the same reference ranges for zinc for both plasma and serum [Wei-jie 1986, Wu 2006]. Comparative studies on nutrient levels in capillary and venous blood have mainly addressed macronutrients like glucose and protein, both of which are 3-10% lower in the venous compared to capillary circulation. Only a few reports have addressed micronutrient levels from different blood draw sites. For example, Kaplan and colleagues measured circulating calcium, potassium, and sodium, but found no statistical difference in capillary versus venous levels [Kaplan et al 1959]. In contrast, Kupke and colleagues found circulating calcium and sodium levels to be lower in capillary blood, but potassium was not different [Kupke et al 1981]. Falch and colleagues also reported circulating calcium and sodium levels to be lower in capillary blood but found potassium to be higher in capillary blood [Falch et al 1981]. Despite these reports, these differences are thought to be of minor clinical importance [Falch et al 1981]. We have found no modern reports that follow up on these studies, which might benefit from the use of more precise methods and technology. Moreover, zinc was not reported in any of these previous studies, so the impact of blood draw site on zinc assessment is not resolved. In this study, we found that capillary blood plasma had an 8% higher mean zinc concentration than venous blood plasma from the same donors in this cohort. This difference is small but shows that different blood draw sites can influence values derived from zinc assessment.

The second pre-analytical variable tested was different blood matrices, specifically plasma and serum formats. Several studies have examined how zinc levels vary between the two matrices but found no difference in zinc content between plasma and serum samples [Kosman 1979, Kiilerich 1981, Keyzer 1983, Chen 1986, Smith 1987, Barroso 2018]. However, other reports did show modest differences in zinc content between plasma and serum samples, with zinc generally higher in serum compared to plasma samples taken from the same individuals [Foley 1968, Kasperek 1981, English 1988], though at least one report found higher zinc in plasma samples [Kraus 2018]. English and Hambidge suggested that the differences in the zinc content of the matrices may be due to the required processing delay to generate serum from whole blood, which could allow more time for zinc to diffuse from cellular sources [English 1988]. Later, more comprehensive analyses of the blood matrix revealed that many nutrients and metabolites were in fact at higher concentration in the serum compared to plasma [Kronenberg 1998, Yu 2011]. These authors proposed that the influence of a volume displacement effect caused by the loss of coagulation proteins led to an increase in the fractional distribution of other metabolites. However, zinc was not measured in those studies, and follow up work to confirm the impact of volume displacement on zinc concentration have yet to be reported. Thus the potential difference in zinc content between plasma or serum remains unresolved, so individual laboratories tend to choose the matrix from their own historical preference, constraints from other co-analytes, or logistical aspects of study design. In fact, much of the commonly cited literature for zinc assessment treat plasma and serum zinc concentrations as equivalent [IZiNCG 2007, Hess 2008, King 2016]. In this study, we found that venous blood serum had a 5% greater mean zinc concentration than venous blood plasma from the same donors in this cohort. This difference is small but shows that different blood matrices can influence values derived from zinc assessment.

The third pre-analytical variable tested was different BCT manufacturers, specifically testing similar BCT types from BD and Sarstedt. We are unaware of any previous study reporting zinc values from different manufacturers, but this should be an important issue since the market share of these two companies differs internationally. The closest matched BCT types had slightly different anticoagulant additives, with BD using K_2_EDTA and Sarstedt using K_3_EDTA. Our goal was to compare manufacturers with similar BCT types, and these products were the closest between the two companies. Publication comparing BCTs with K_2_EDTA and K_3_EDTA for several analytical targets showed minimal differences between the anticoagulants, though zinc was not specifically measured [Brunson 1995, Van Cott 2003]. In this study, the mean zinc concentration for venous blood plasma or serum collected in BD BCTs was not different between the two manufacturers. However, the mean zinc concentration for capillary blood plasma collected in BD BCTs was 6% higher than capillary blood plasma collected in Sarstedt BCTs from the same donors. One explanation for this could be that the BD capillary BCTs had a higher zinc background than the Sarstedt capillary BCTs. Neither BD nor Sarstedt capillary BCTs were certified for use with zinc assessment, but a random sampling of empty tubes from the study lot numbers did not indicate any significant contamination of zinc from either manufacturer. It is important to note that only a single lot number from each manufacturer was used in this study, so it is not possible to state that one manufacturer was superior to another. However, the implication is that even the type of BCTs used can affect zinc assessment, so testing before use and avoiding tube type mixing is highly recommended.

The fourth and fifth pre-analytical variables were time until processing and holding temperature for blood samples, respectively. These parameters are often examined together given their interrelationship. Several studies have described changes in nutrient levels as a function of processing time and temperature [Hankinson 1989, Key 1996, Drammeh 2008, Cray 2009, Cuhadar 2013], but few of these included zinc as an analyte. One of the first reports on zinc was by English and Hambidge who found that both plasma and serum zinc increased by 5% when whole blood processing was delayed by 2 hours, though the samples were kept at ‘room temperature’ [English 1988]. Later studies corroborated these findings by showing significant increases in plasma zinc when samples were held at ‘room temperature’ for 2-4 hours [Kraus 2016, Chovelon 2018, Leger 2020]. An early comparison of both processing time and temperature by Tamura and colleagues found that plasma and serum zinc were significantly increased between 1-5 hours when whole blood samples were kept at 23°C, but replicate samples kept at 7°C were not significantly increased until 24 hours [Tamura 1994]. An important caveat is that all previously cited studies used heparin as the anticoagulant for plasma samples unlike the current study that used EDTA, though at least one report showed no difference between EDTA and heparin for determining zinc levels in plasma [Bao 2021]. However, we are not aware of any reports that directly compare zinc from serum samples with plasma samples using EDTA. We also could not find reports in which zinc was measured from blood kept at warmer temperatures, like what might occur in an equatorial country without immediate access to a cold chain. Studies looking at other metabolites did show significant changes when plasma was kept at 37°C for even 1 hour [Yin 2015]. In this study, the mean zinc concentration for venous blood plasma and serum ranged from 5-12% when processing was delayed 4 or 24 hours after collection. Additionally, the mean zinc concentration for venous blood plasma and serum ranged from 0.5-7% when incubated at 20°C or 37°C after collection. These results are consistent with previous work showing that both time until processing and holding temperature can have a significant influence on zinc assessment.

A strength of this study is that important pre-analytical variables were evaluated under a single chain of custody with tight control over most of the other environmental and technical parameters that could add to zinc measurement variance. These results help fill in literature gaps on the impact of blood draw site, tube types, and holding procedures during zinc assessment. Yet the highly controlled structure of this study is also a limitation, as the control obtained with the clinical laboratory does not reflect the practical challenges of conducting a large cohort or field assessment of zinc status. However, it seems important to understand the baseline variability within these pre-analytic variables in order to properly calculate how study factors will affect the dispersion of zinc measurements. Another limitation of this study is that the plasma and serum samples held for 4 or 24 hours for later processing were frozen and analyzed on a different day from the time 0-hour samples due to logistical reasons. The ICP was calibrated with the same standards and validated with the same quality control samples on each run day, but we cannot rule out that the freeze/thaw process or instrument performance differences could have influenced zinc measurements. An additional limitation is that not all BCTs or other clinical supplies used in this study were available as trace metal-certified products. We sampled all of the BCTs and other supply lots for zinc contamination before use but cannot guarantee every consumable was zinc free. The difficulties in matching the preferred BCT to the study design illustrates the need for more trace metal-certified options for clinical assessments of circulating zinc.

This study attempted to quantify the importance of specific pre-analytical variables on the measurement of circulating zinc concentrations, which is key to determining the public health burden of zinc deficiency in target populations. Blood draw site, blood matrix, blood collection tube type, blood processing delay, and blood holding temperature all had small but significant impacts on the assessment of circulating zinc levels. Thus, caution is warranted when comparing studies that use different procedural methods to measure zinc concentration in plasma or serum.

Furthermore, future studies should endeavor to control these pre-analytical variables in order to return the highest quality data for zinc assessment.

## Data Availability

All data produced in the present study are available upon reasonable request to the authors.

## Acknowledgements

The authors thank Nyla Sepulveda for expert phlebotomy service during this study. The authors also thank Bonny Alvarenga for laboratory assistance, Dr. Ellen Fung for providing clinical space, Maria Cruz for assistance with supplies, and Sarstedt, Inc. for donating blood collection tubes. Additionally, the authors thank Dr. Christine McDonald, Dr. Mari Manger, and the IZiNCG Steering Committee for helpful feedback.

---

**S1 Table.** Demographic data from study participants. Self-identified demographic data are shown for participants in this study. For demographic data, all participants selected a female or male identifier; no participant selected nonbinary or other choice. Body-mass index values are indicated in ‘BMI’ column. The categorical abbreviations include “AA” for African American, “Latinx” for a gender-neutral indication of Latin American origin, and “Multiple” for multiple race/ethnicity identity.

| participant # | age range | gender | race/ethnicity | supplement use | BMI |
| --- | --- | --- | --- | --- | --- |
| 1 | 40-49 | female | White | occasional | 19.0 |
| 2 | 40-49 | male | White | never | 41.0 |
| 3 | 50-59 | female | White | frequently | 19.6 |
| 4 | 50-59 | female | White | frequently | 33.6 |
| 5 | 30-39 | female | White | daily | 30.6 |
| 6 | 18-29 | female | Asian | rarely | 27.4 |
| 7 | 60-69 | male | White | occasional | 31.5 |
| 8 | 18-29 | female | Latinx | occasional | 20.0 |
| 9 | 50-59 | male | AA/Black | daily | 32.7 |
| 10 | 50-59 | female | AA/Black | frequently | 19.3 |
| 11 | 30-39 | female | White | never | 21.2 |
| 12 | 40-49 | male | Latinx | rarely | 21.4 |
| 13 | 60-69 | female | White | occasionally | 23.7 |
| 14 | 18-29 | male | AA/Black | never | 31.1 |
| 15 | 30-39 | male | Asian | never | 22.5 |
| 16 | 40-49 | female | White | rarely | 29.1 |
| 17 | 50-59 | male | White | occasionally | 23.5 |
| 18 | 18-29 | female | Multiple | never | 24.4 |
| 19 | 30-39 | male | Asian | never | 22.0 |
| 20 | 18-29 | male | Latinx | daily | 29.3 |
| 21 | 18-29 | female | White | never | 27.3 |
| 22 | 60-69 | male | White | occasionally | 24.8 |
| 23 | 40-49 | female | Latinx | never | 26.1 |
| 24 | 30-39 | male | White | never | 20.9 |
| 25 | 50-59 | female | AA/Black | rarely | 24.1 |
| 26 | 50-59 | female | Asian | daily | 22.2 |
| 27 | 18-29 | male | Multiple | frequently | 39.5 |
| 28 | 50-59 | male | Asian | occasionally | 25.1 |
| 29 | 40-49 | male | White | rarely | 25.3 |
| 30 | 30-39 | female | Asian | rarely | 29.3 |
| 31 | 30-39 | female | Asian | rarely | 21.6 |
| 32 | 60-69 | female | Multiple | daily | 44.4 |
| 33 | 50-59 | male | Asian | never | 30.4 |
| 34 | 60-69 | male | White | daily | 23.6 |
| 35 | 40-49 | female | Multiple | occasionally | 26.1 |
| 36 | 18-29 | male | Latinx | frequently | 25.8 |
| 37 | 30-39 | male | AA/Black | never | 26.9 |
| 38 | 18-29 | female | Multiple | daily | 21.5 |
| 39 | 18-29 | male | Asian | never | 21.8 |
| 40 | 60-69 | male | White | occasionally | 25.0 |
| 41 | 18-29 | female | Asian | rarely | 33.1 |
| 42 | 18-29 | male | Multiple | rarely | 31.8 |
| 43 | 18-29 | female | White | never | 20.7 |
| 44 | 18-29 | female | Asian | daily | 17.1 |
| 45 | 18-29 | male | Latinx | occasionally | 22.1 |
| 46 | 18-29 | female | Latinx | rarely | 32.7 |
| 47 | 30-39 | male | Asian | never | 25.4 |
| 48 | 60-69 | female | White | occasionally | 34.3 |
| 49 | 18-29 | female | White | never | 21.0 |
| 50 | 30-39 | female | Asian | occasionally | 19.4 |
| 51 | 18-29 | male | White | occasionally | 25.3 |
| 52 | 40-49 | female | AA/Black | never | 23.4 |
| 53 | 40-49 | female | White | frequently | 27.4 |
| 54 | 40-49 | female | Middle Eastern | daily | 20.7 |
| 55 | 30-39 | male | Asian | occasionally | 25.5 |
| 56 | 18-29 | female | Asian | occasionally | 22.0 |
| 57 | 60-69 | male | White | daily | 34.3 |
| 58 | 18-29 | female | Asian | rarely | 22.1 |
| 59 | 60-69 | male | White | daily | 27.1 |
| 60 | 40-49 | female | White | daily | 29.2 |

**S2 Table.**
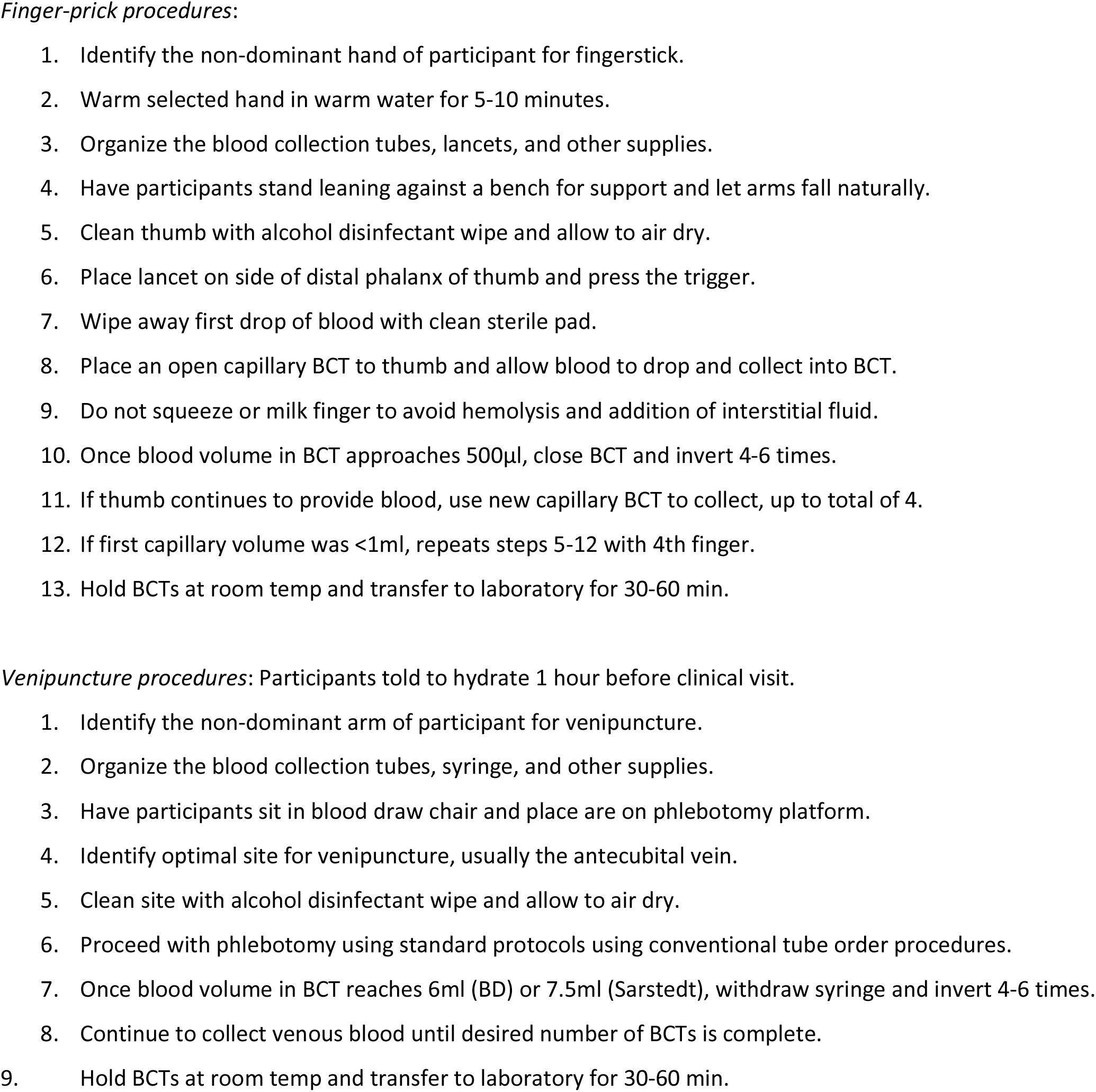
The procedure for sequential collection of venous and capillary blood samples for this study are listed. “BCT” indicates blood collection tubes.

**S3 Table.**
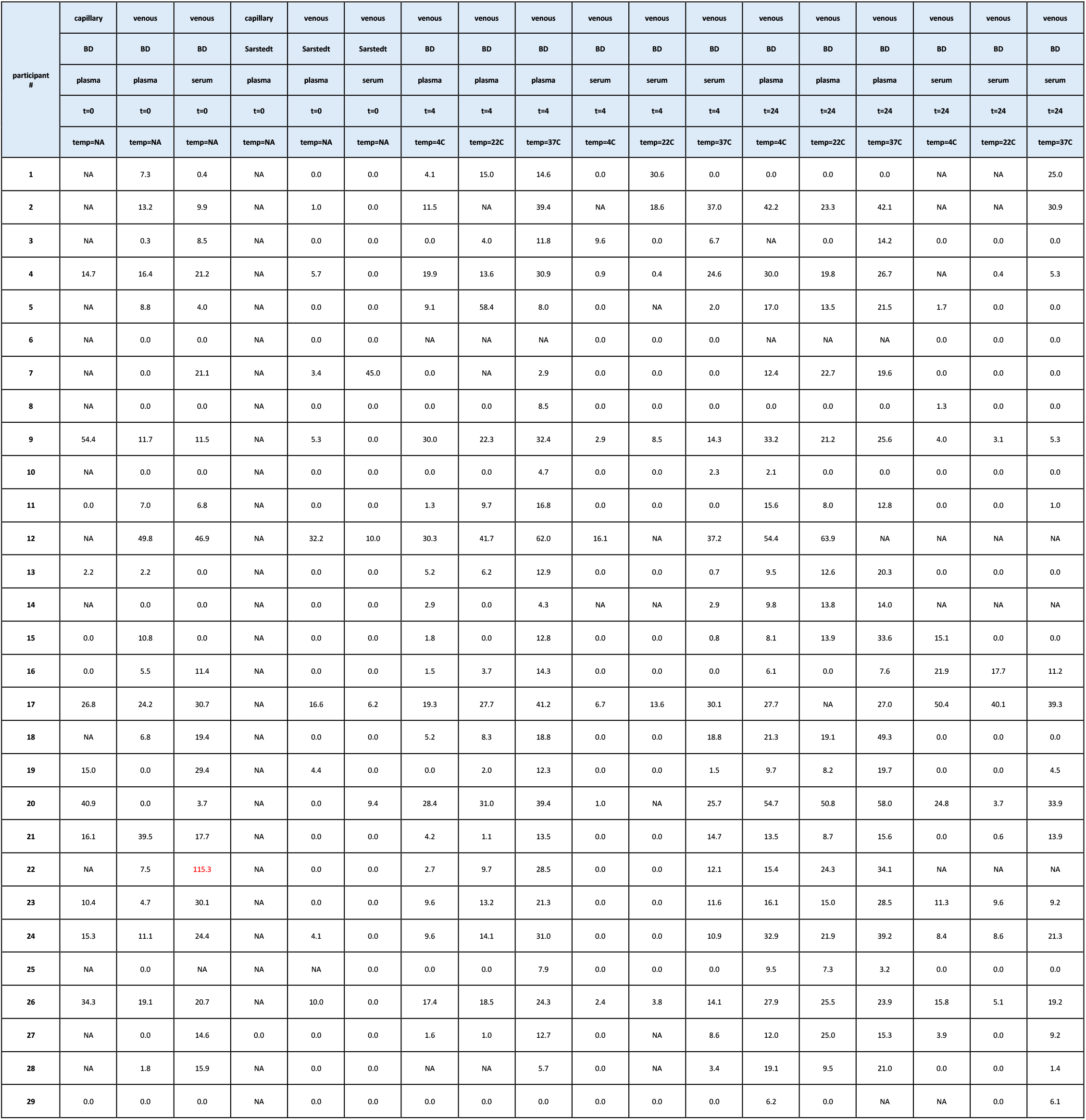
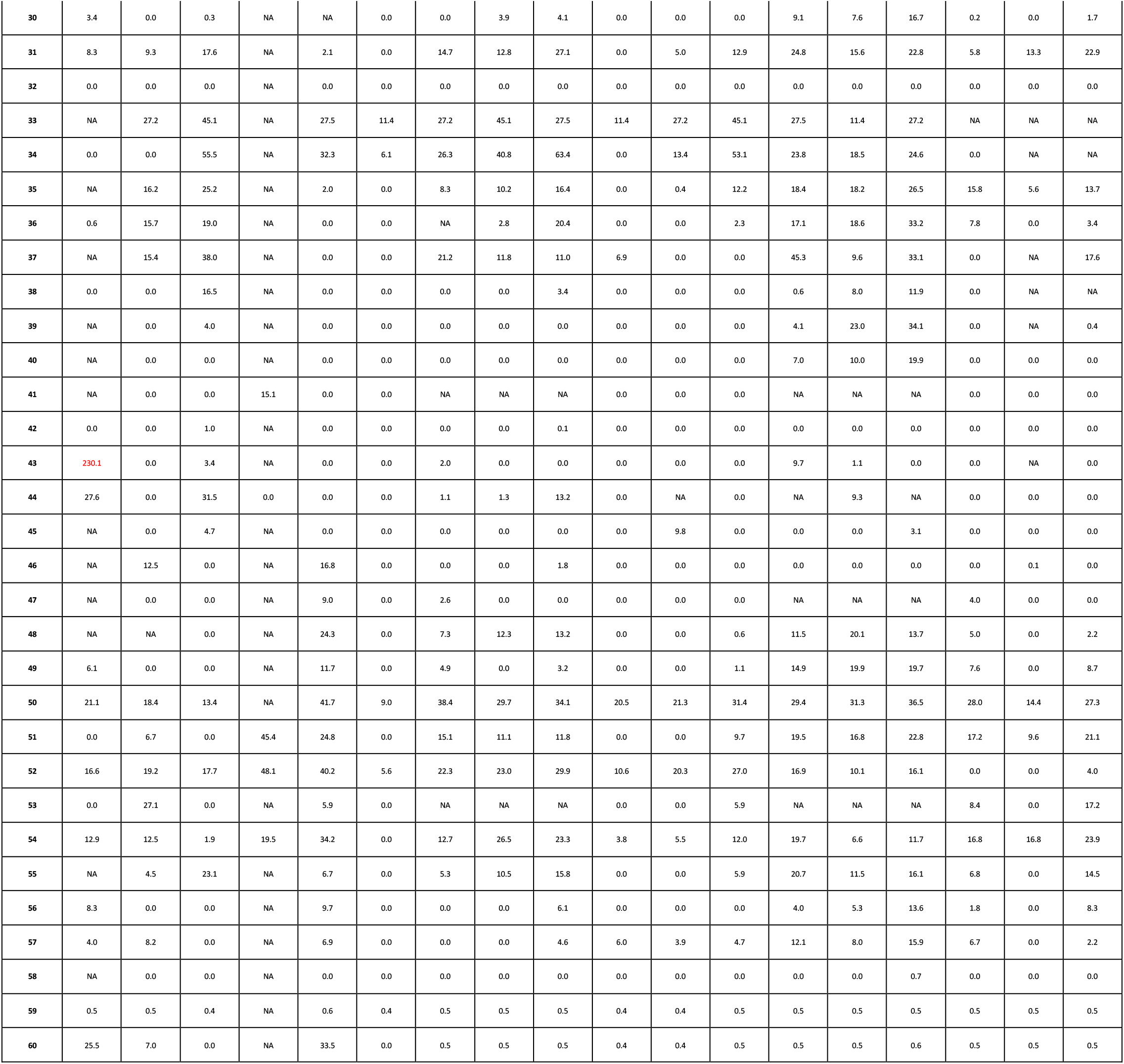
Hemolysis levels in all study participant samples. Hemolysis is indicated by hemoglobin content (mean in mg/dL, *n*=1-3) for all plasma or serum samples at all blood draw sites, blood matrices, blood collection tubes manufacturers, processing delay, and holding temperatures are shown. Samples processed immediately are marked t=0 and temp=NA. Processing delay is indicated for 4 hour (t=4) or 24 hours (t=24). Holding temperature is indicated for 4°C (temp=4C), 20°C (temp=20C), or 37°C (temp=37C). Missing values due to lack of blood volume are identified with an ‘NA’. Hemoglobin values greater than 100 mg/dL (elevated hemolysis) are identified with red font.

**S4 Table.**
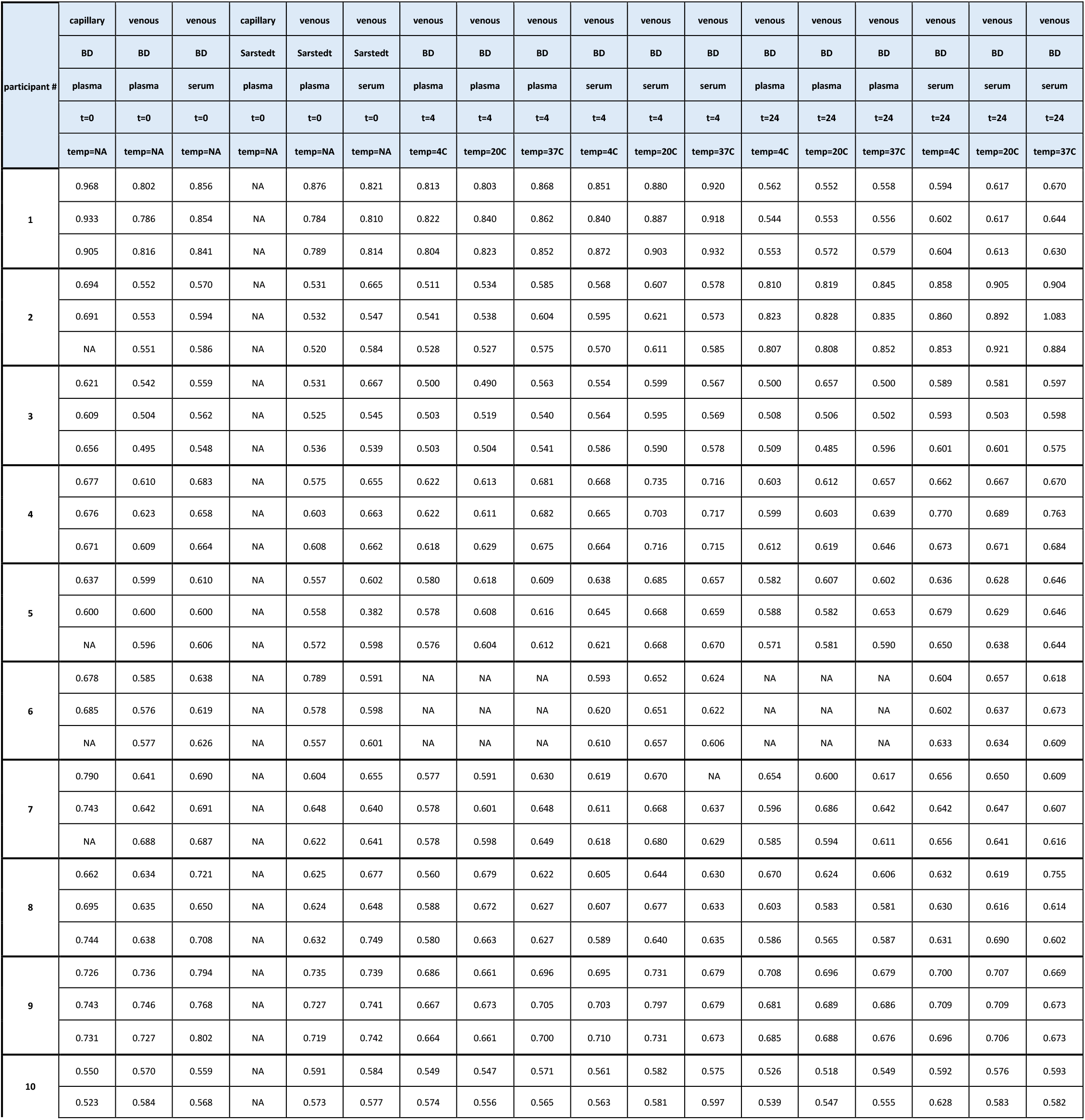
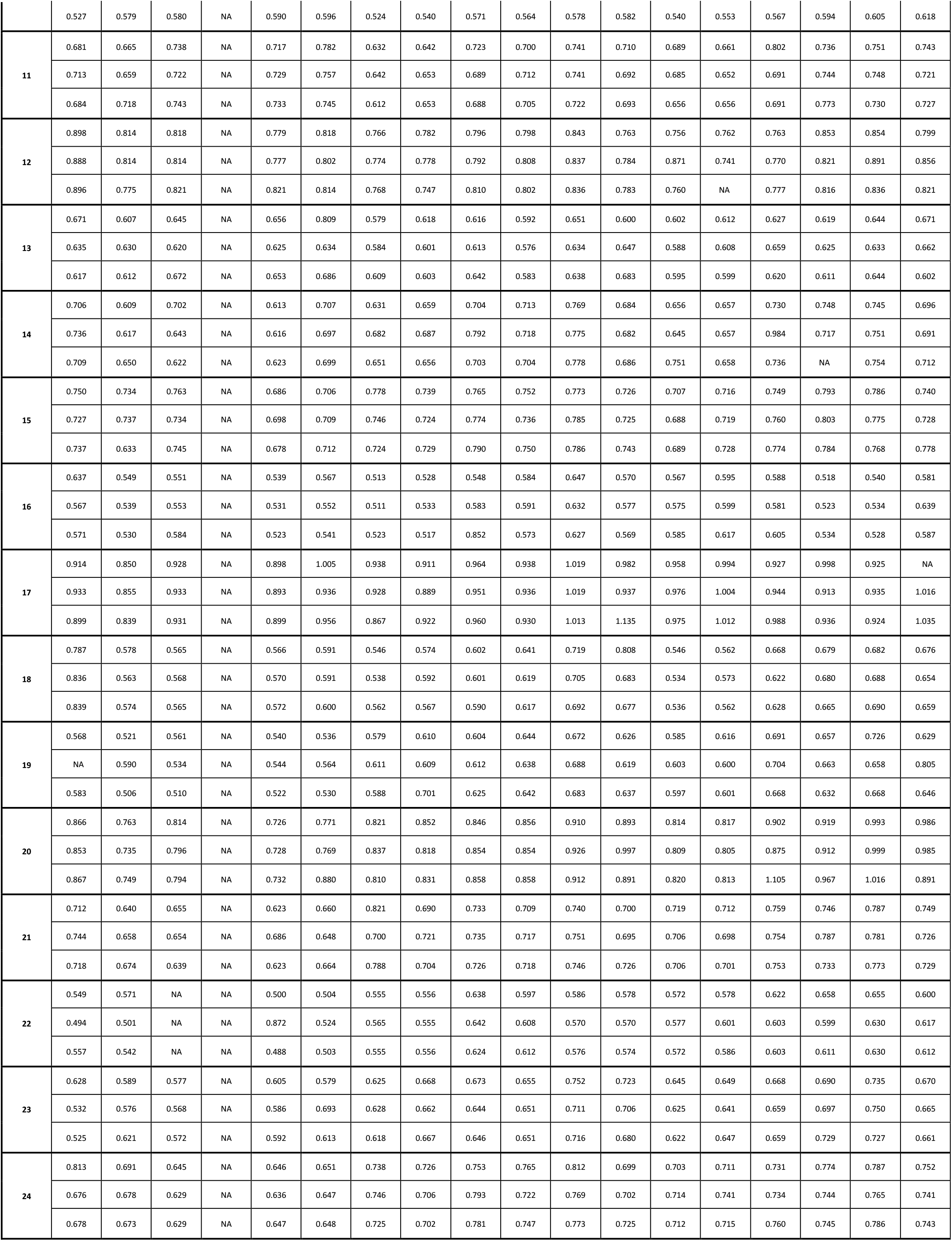
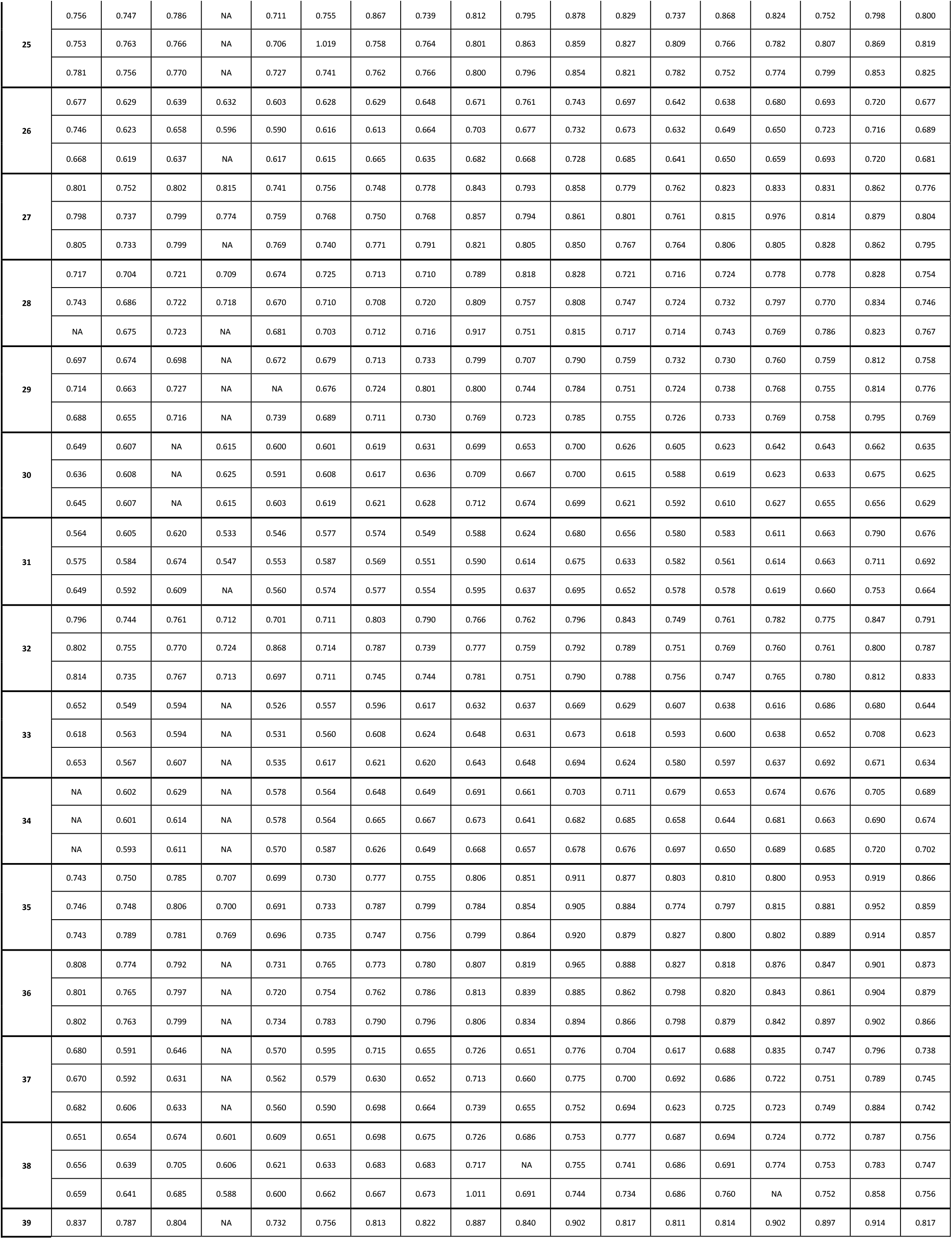
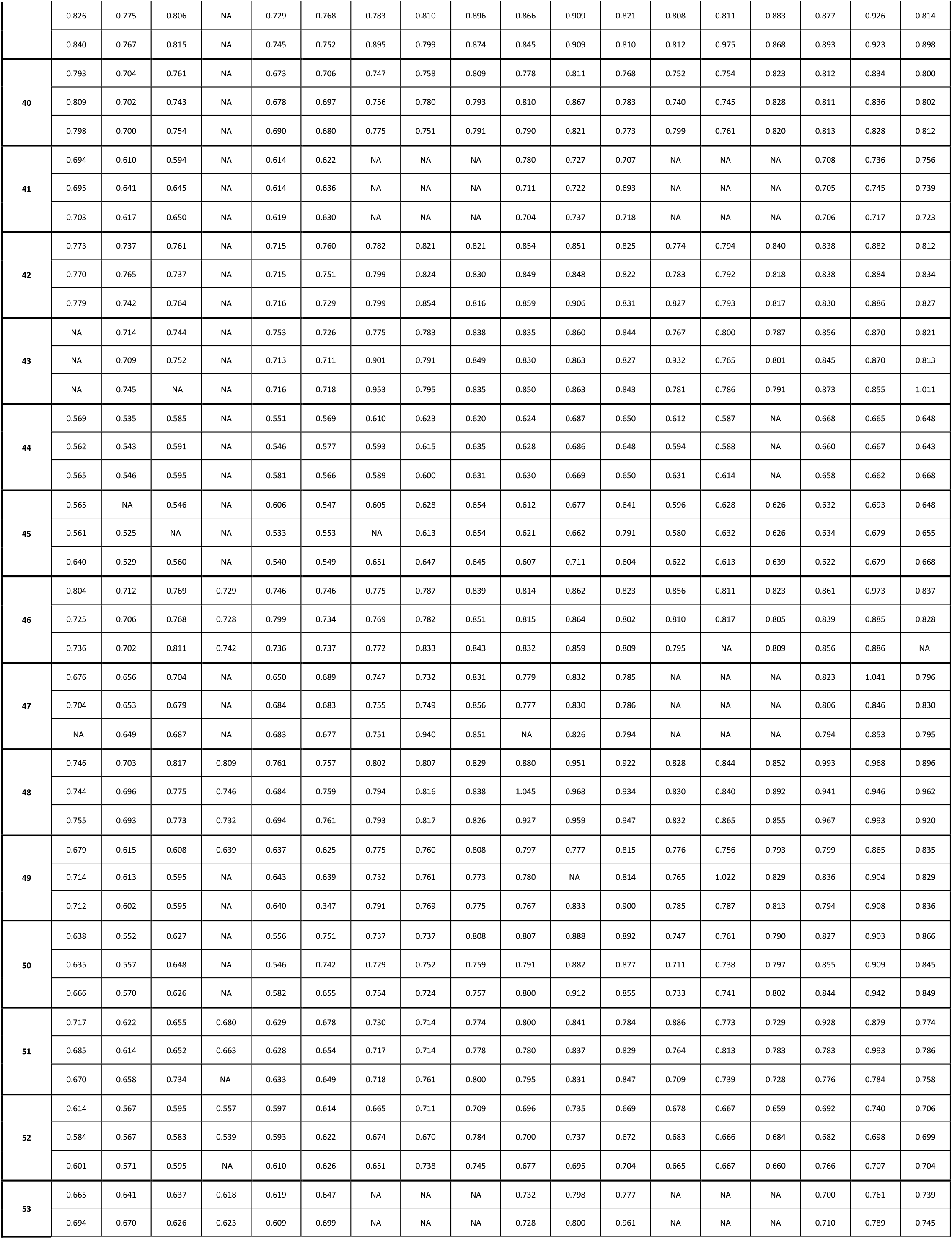
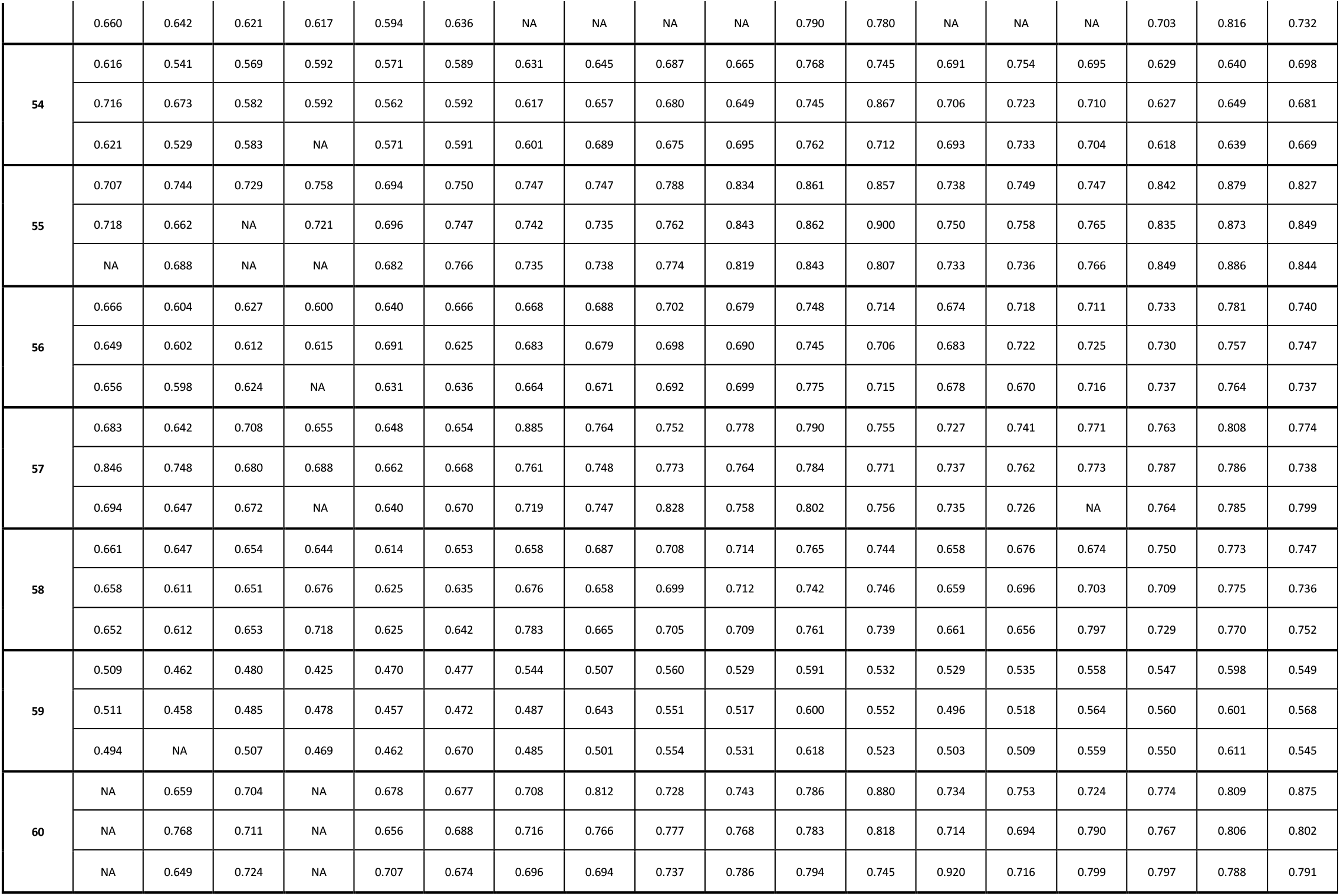
Zinc values in all study participant samples. The zinc measurements for participants for all blood draw sites, blood matrices, blood collection tubes manufacturers, processing delay, and holding temperatures are shown. Triplicate values (in mg/L) are shown for all plasma or serum samples. Samples processed immediately are marked t=0 and temp=NA. Processing delay is indicated for 4 hour (t=4) or 24 hours (t=24). Holding temperature is indicated for 4°C (temp=4C), 20°C (temp=20C), or 37°C (temp=37C). Missing values due to lack of blood volume are identified with an ‘NA’.

**S1 Fig:**
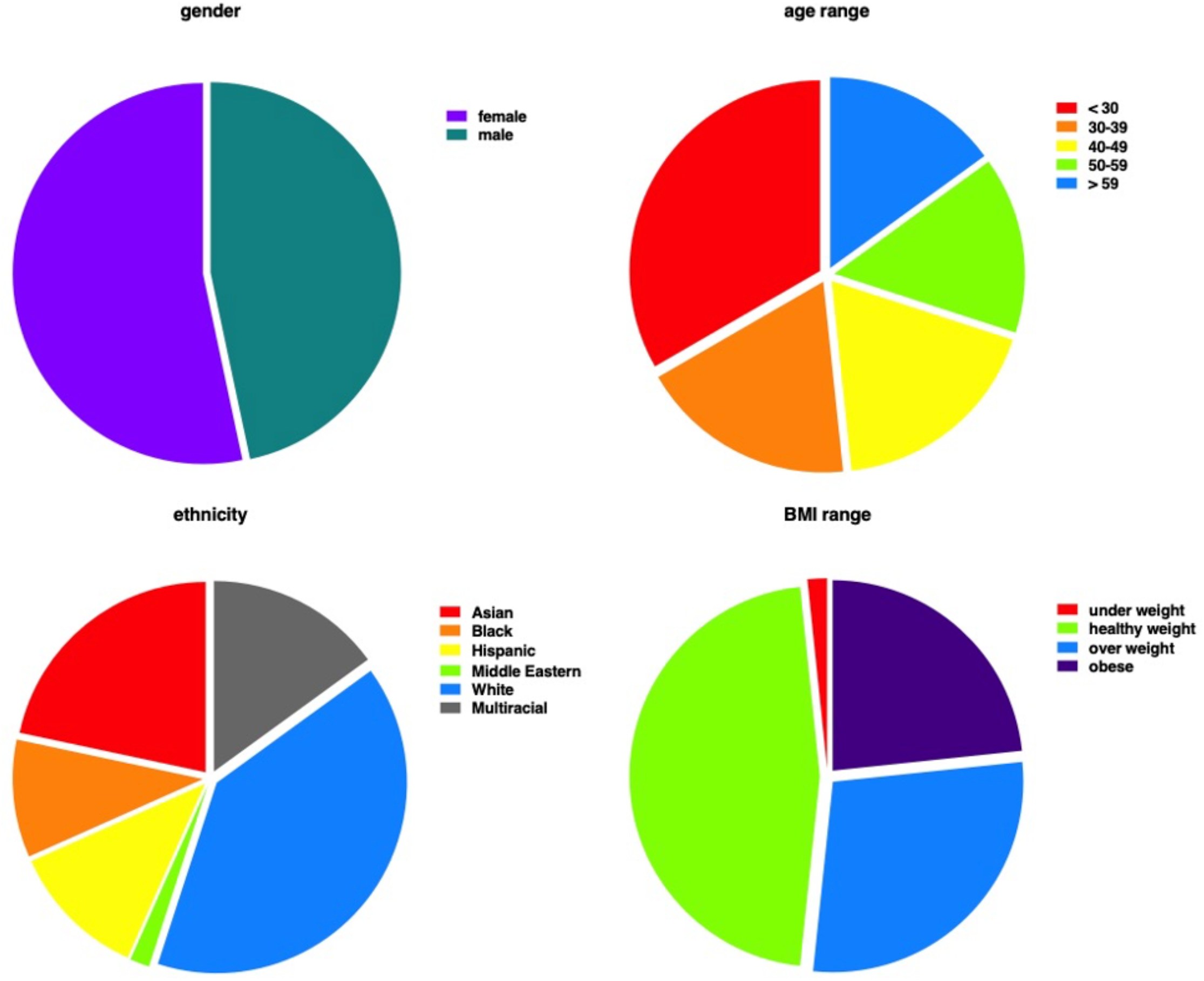
Summary of demographic information on study participants. Charts show gender, age-range, race/ethnicity, and body mass index (BMI) for participants. Gender, age, and ethnicity were provided directly by participants. BMI was calculated from height and weight measured on each participant.

**S2 Fig:**
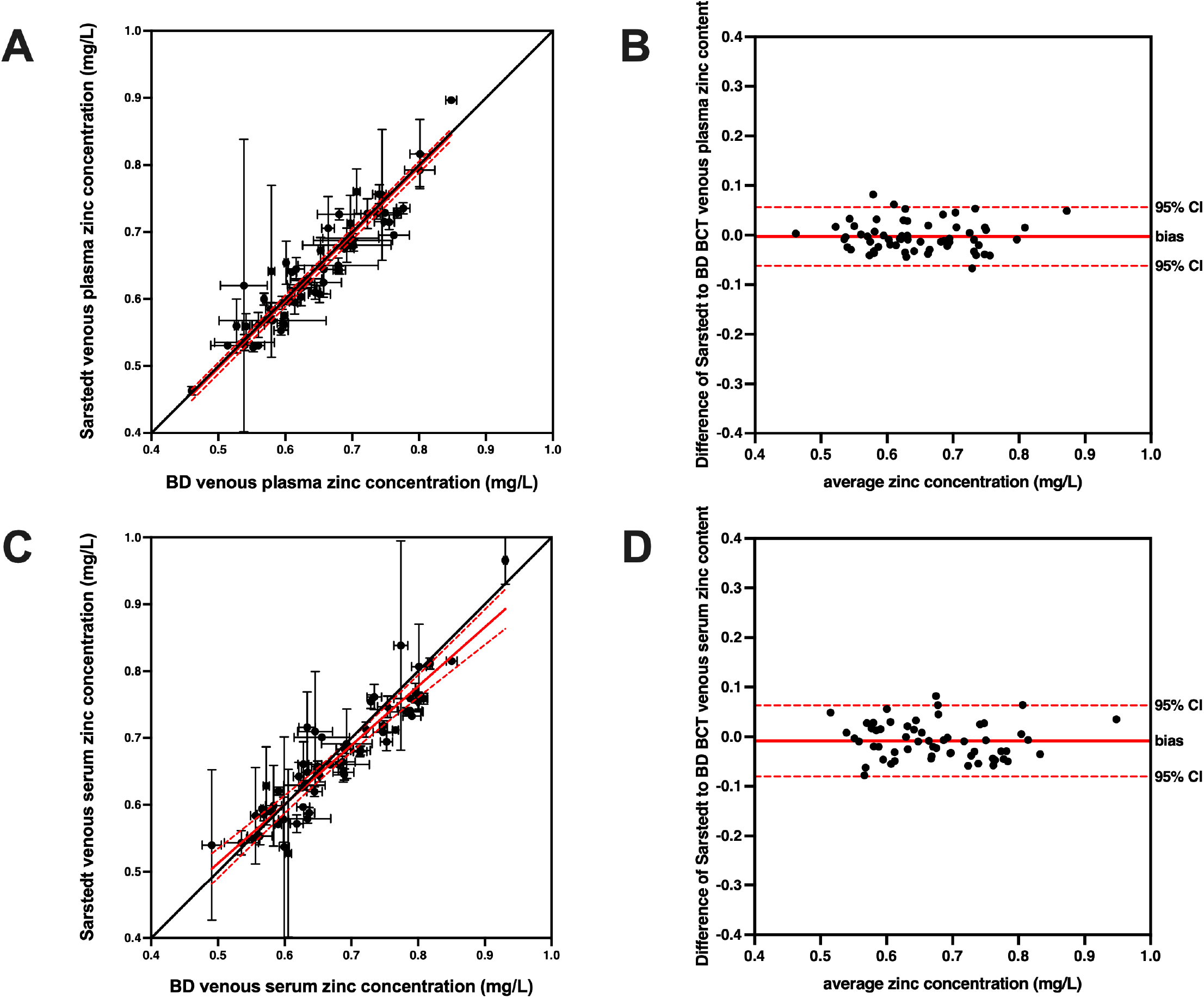
Zinc concentrations in venous samples do not differ by blood collection tube (BCT) manufacturer. Correlation plots comparing circulating zinc values from **(A)** venous plasma from BD and Sarstedt BCTs and **(C)** venous serum from BD and Sarstedt BCTs are shown. Each circle represents the zinc level (mean ± SD, *n*=1-3) for an individual participant, with linear regression and 95% confidence interval indicated by a solid red and dotted red line, respectively. The line of concordance is shown as a solid black line for comparison. Since there was no significant difference in the mean zinc concentrations for either matrix, Bland-Altman plots are only shown for illustrative purposes for **(B)** venous plasma and **(D)** venous serum samples. No significant bias was measured. Each circle represents the zinc level for an individual participant, with average distance and 95% confidence interval indicated by a solid red and dotted red line, respectively.

